# Environmental Exposure to Respirable Particles and Estimated Risk of Sarcoidosis: A Systematic Review and Meta-Analysis

**DOI:** 10.1101/2022.04.19.22274019

**Authors:** David Wambui, Ogugua Ndili Obi, Guy Iverson, Kevin O’Brien, Gregory Kearney

## Abstract

**Introduction:** Sarcoidosis is an inflammatory disease of unknown etiology that affects multiple organs in the body. In most cases, the affected organ is the lung. Sarcoidosis risk factors include environmental exposures, genetic predisposition, and immunological factors. The main objective of this review was to assess whether exposure to respirable particles is associated with increased risk of sarcoidosis.

**Methods:** A comprehensive search was conducted in scientific databases. Additional search of grey literature as well as handsearching of relevant records was performed. The search was restricted to studies published between January 1998 to October 2019. Meta-analysis was performed for studies that provided quantitative data.

**Results:** After applying inclusion/exclusion criteria, nine articles were included in the systematic review and four in the meta-analysis. Quantitative analysis suggested that people exposed to respirable particles were approximately three times more likely to develop sarcoidosis compared to people who are unexposed.

**Discussion and conclusion:** This study collected and aggregated available evidence that assessed exposure to respirable particles and risk of developing sarcoidosis. Evidence of increased association between exposure to respirable particles and sarcoidosis was strongly suggested based on our qualitative review. More rigorous epidemiologic exposure studies are needed to generate data that would accurately determine the risk and causal pathways through which exposure to respirable particles could lead to the development of sarcoidosis.

## Introduction

Sarcoidosis is an inflammatory multisystem disease that presents with noncaseating granulomas in affected organs (1). The most common organ affected in over 90% of cases is the lungs (2). The disease has no known etiology but has been associated with several risk factors including occupational and environmental exposures (3). In a study conducted in Sweden among iron foundry workers, exposure to silica dust was significantly associated with increased risk of sarcoidosis (4). In similar studies, it was observed that cessation of exposure to silica resulted in regression of sarcoid granulomas (5,6). In Iceland, the incidence rate of sarcoidosis among limestone workers was up to 4 times higher than that of the general population (7). However, a study conducted in Italy to investigate the probable correlation between environmental exposures including mineral particle air pollution failed to conclusively determine any association between environmental exposure and risk of sarcoidosis (8).

In the US, several studies have shown an increased incidence of sarcoidosis among emergency responders and firefighters (9–11). Webber and colleagues (12) found an increased incidence (25 per 100,000) of sarcoidosis among male rescue/recovery workers 12 years after the World Trade Center (WTC) bombings on September 11, 2001. It has been hypothesized that occupational exposures to varying products and substances, such as particulate matter (PM), fibers, volatile organic pollutants, gases, metal dusts and other combustion byproducts from the WTC fires could be considered etiological factors that trigger inflammation or an abnormal immune response in the body that lead to the development of sarcoidosis (13). Although several studies provide convincing evidence of an association between exposure to respirable particles and sarcoidosis, there has not yet been a systematic review and meta-analysis published aggregating the evidence.

The objectives of this study were to 1) critically evaluate the literature related to environmental or occupational exposures of respirable PM, including dust, and the association with physician diagnosed sarcoidosis, and 2) conduct a meta-analysis to assess the strength of measured effects found from relevant studies. The results of this study will provide a risk estimate of the effect of environmental/occupational exposure to respirable PM and sarcoidosis.

## Methods

### Search Strategy, Inclusion/Exclusion Criteria

The Preferred Reporting Items for Systematic Reviews and Meta-Analysis (PRISMA) Statement (14) was used to critically appraise and synthesize the published literature. Scientific database search engines were used to identify relevant articles using keywords, titles, and abstracts (Table 1). The search was restricted to 1) articles published in English, 2) observational studies (i.e., cohort, case analysis/case series, and case-control studies), and 3) studies published between January 1, 1998, to October 31, 2019. The following medical subject headings (MeSH) terms were used to search the electronic databases; (respirable particles OR dust OR particulate matter) AND (sarcoidosis). Hand-searching of grey literature (government and institutional reports) and reviewing relevant references from published articles was conducted to identify additional studies.

**Table 1.** Databases searched for articles published between 1998 and 2019

| Search Engine Database |
| --- |
| 1. PubMed |
| 2. Ovid |
| Medline |
| Embase and Embase Classic |
| 3. Scopus |
| 4. ProQuest Central |
| 5. Google Scholar |
| 6. Cochrane Library |

### Data Synthesis

Eligibility criteria for a paper to be included in the meta-analysis were 1) a clearly defined measure of associated risk (i.e., relative risk ratio or odds ratio; 2) ICD 10 classification (D86.0 - sarcoidosis of the lung; D86.1 - sarcoidosis of lymph nodes; D86.2 - sarcoidosis of the lung with sarcoidosis of lymph nodes; D86.3 - sarcoidosis of skin; D86.8 - sarcoidosis of other and combined sites; D86.9 - unspecified sarcoidosis) or noted as a physician-diagnosed sarcoidosis case, and 3) observational studies which included an exposure group, and/or individuals who had prior exposure to respirable dust particles. For the last criterion, studies had to include a comparison or control group with no prior exposure to respirable dust particles. Case-control studies needed to include matched comparisons (e.g., gender, age, occupation) as applicable to eliminate potential sampling and/or selection bias. Respirable dust particles were defined as particles with an aerodynamic diameter of 10 micrometers (µm) or less in size (PM_10_ or lesser).

Following a comprehensive literature search and retrieval of all publications that met the inclusion criteria, a thorough review was conducted by two independent reviewers (DW and GK). Studies that included quantitative data were included in the meta-analysis. Any differences in the review process were resolved through consensus.

### Data Extraction

Information and data from the search were conducted in accordance with the PRISMA statement guidelines and included article title, authors’ information, sample size, study population, follow-up time, outcome measure, and estimated risk (risk ratio or odds ratio) where applicable. This study involved secondary data analysis was considered exempt for approval by the Institutional Review Board at East Carolina University.

### Data analysis

A meta-analysis was conducted using RevMan V5.3 software from the Cochrane Collaboration (London, UK). The numbers of cases and controls provided by individual studies were entered into RevMan and pooled risk estimates were recalculated using a random effect model to generate study weights. Weights for individual studies were used to compute a pooled risk estimate. To test the heterogeneity of the studies included, I^2^ statistic was calculated. With only few studies eligible for this systematic review and meta-analysis, reporting bias was minimal. Searching for grey literature also minimized the risk of publication bias.

## Results

As shown in Figure 1, the initial literature search yielded 105 articles. Auxiliary searches identified from citations and references in key papers yielded an additional three articles for a total of 108 articles. After removing duplicate articles, 66 records remained. After screening titles and abstracts, it was determined that 56 articles did not meet the inclusion criteria, leaving 13 records that went through full review. Four studies were excluded after full review since they did not assess risk. Among those nine articles that met the eligibility criteria, only four had quantitative data and were therefore included in the meta-analysis. A summary of studies included in the review is shown in Table 2.

**Figure 1.**
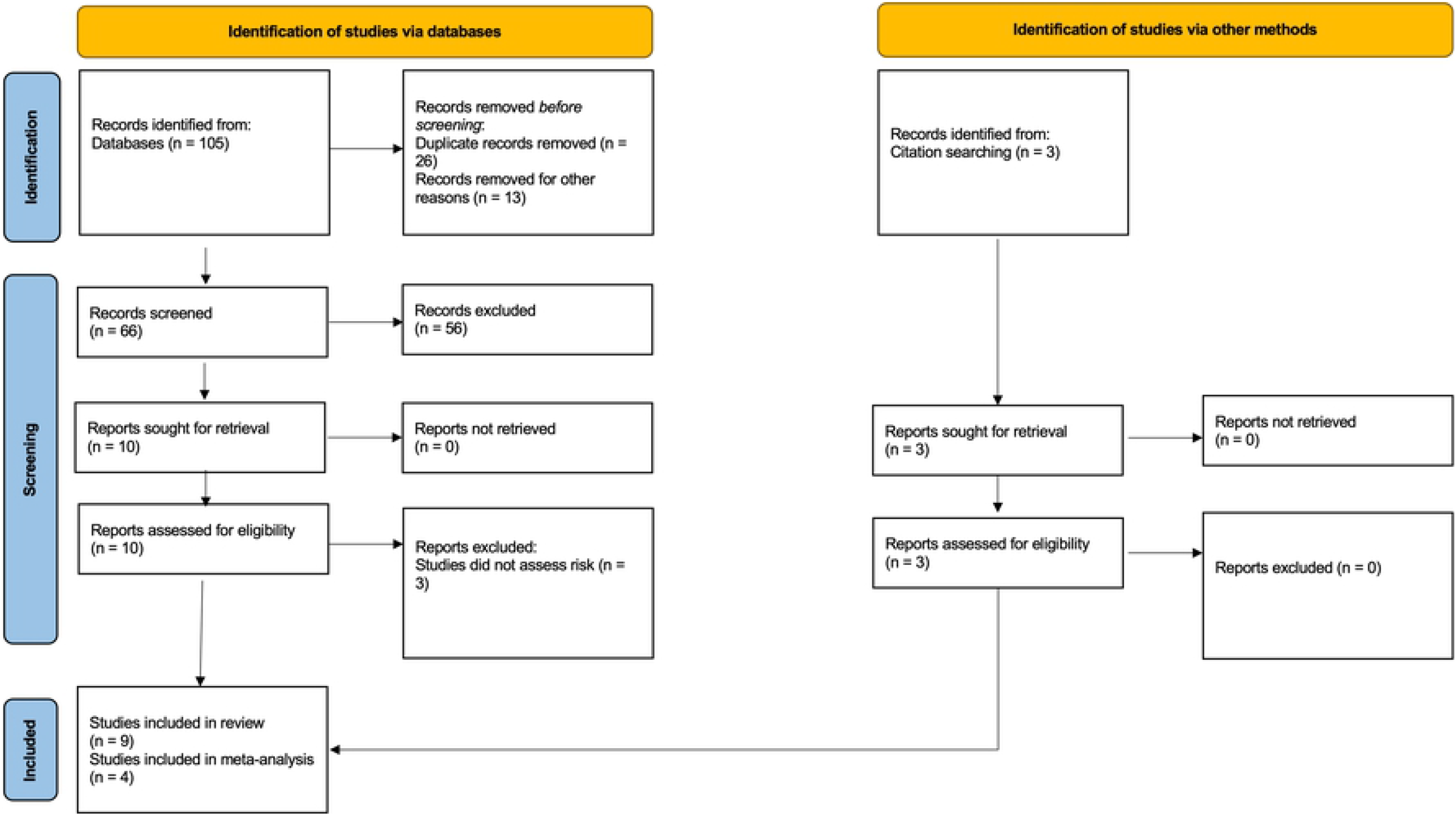
PRISMA Flow Diagram (14)

**Table 2.** Summary results of eligible sarcoidosis and exposure studies published between 1998-2019

| Study author(s) | Study design | Publication year | Case definition | Year(s) of study | Follow-up period (years) | No. of Cases | No. of Controls | Sample size | Mean age | Participant/ occupation description | Country | Relative Risk (95% CI) |
| --- | --- | --- | --- | --- | --- | --- | --- | --- | --- | --- | --- | --- |
| Vihlborg et al. (4) | Retrospective Cohort | 2017 | ICD-10 (D86-0-3,8); RA M05 (0-9) and M06 (0-4,8,9) 9 code 135) | 2001-2013 | 12 | 7 | - | 2187 | - | Iron foundry workers | SE | 3.92 |
| Hena et al. (16) | Retrospective cohort | 2019 | ICD-10 (D86) | 2005 - 2018 | 13 | 87 | 5144 | 11600 | 41.5 | WTC Bellevue hospital | USA | 0.90 |
| Sunil et al. (17) | Case evaluation | 2019 | ICD-10 (D86) | 09/2001 - 12/2001 | - | 6 | - | - | - | WTC cohort | USA | NP |
| Crowley et al. (18) | Case evaluation | 2011 | (ICD-9 code 135) ;(ICD-9 code 516.3 | 2008 | - | 38 | - | 19756 | - | WTC responders | USA | NP |
| Izbicki et al. (19) | Case analysis | 2007 | ICD-10 (D86) | 2001-2006 | 5 | 26 | - | - | - | WTC responders | USA | NP |
| Hena et al. (20) | Retrospective cohort | 2018 | ICD-10 (D86) | 2001 - 2015 | 14 | 59 | - | - | - | WTC firefighters | USA | NP |
| Pirozzi et al. (21) | Cohort | 2018 | ICD-10 (D86) | 2013 - 2015 | 2 | 16 | - | - | 59 | University of Cincinnati Medical Center | USA | NP |
| Rafnsson et al. (7) | Case-Control | 1998 | - | 1974 - 1993 | 19 | 8 | 70 | 4500 | - | Husavik heath center/hospital | Iceland | 13.2 |
| Jordan et al. (10) | Case-Control | 2011 | Tissue biopsy (noncaseating granulomas) | 2002 - 2008 | 6 | 43 | 45899 | 45942 | - | WTC health registry | USA | 1.0 |

Note: WTC – World Trade Center; NP – not provided

Out of the nine studies reviewed, six evaluated the exposure among the 9/11 WTC responders or firefighters and their risk of sarcoidosis. Three out of the six studies that focused on WTC cohort were case series (17–19). Two studies were specific to the chemical composition of particulate exposure that they assessed which was silica dust (4,7). One cohort study assessed the association between exposure to particulate air pollution and severity of pulmonary and quality of life symptoms (21). The studies included in the review demonstrated biological plausibility of development of granulomas following exposure to respirable particles, worsening of symptoms, and increased incidence and prevalence of the disease.

Quantitative analysis of evidence in four papers that met the inclusion criteria for a meta-analysis was inconclusive on the risk of development of sarcoidosis due to environmental or occupational exposure to dust. The recalculated risk estimates were slightly different from those provided in individual studies because of the weight and estimation method used. The summarized results for the meta-analysis are shown in Figure 2. The subgroup analysis indicates that studies among European populations indicated significantly higher odds (odds ratio = 15.3, 95% CI, 3.37 - 69.52) of sarcoidosis among the exposed group. The odds ratio from the two studies utilizing the WTC cohorts were not statistically significant as well as the overall odds ratio (odds ratio = 2.7, 95% CI, 0.62 – 11.77). Three of the four studies indicated higher odds of developing sarcoidosis among those exposed compared to the unexposed (4,7,10). However, only one study showed statistically significant higher odds among those exposed (7). Two of the studies included in the meta-analysis assessed the risk of developing sarcoidosis among those exposed to silica respirable particles (4,7). These findings were consistent with the findings of the individual studies included.

**Figure 2.**
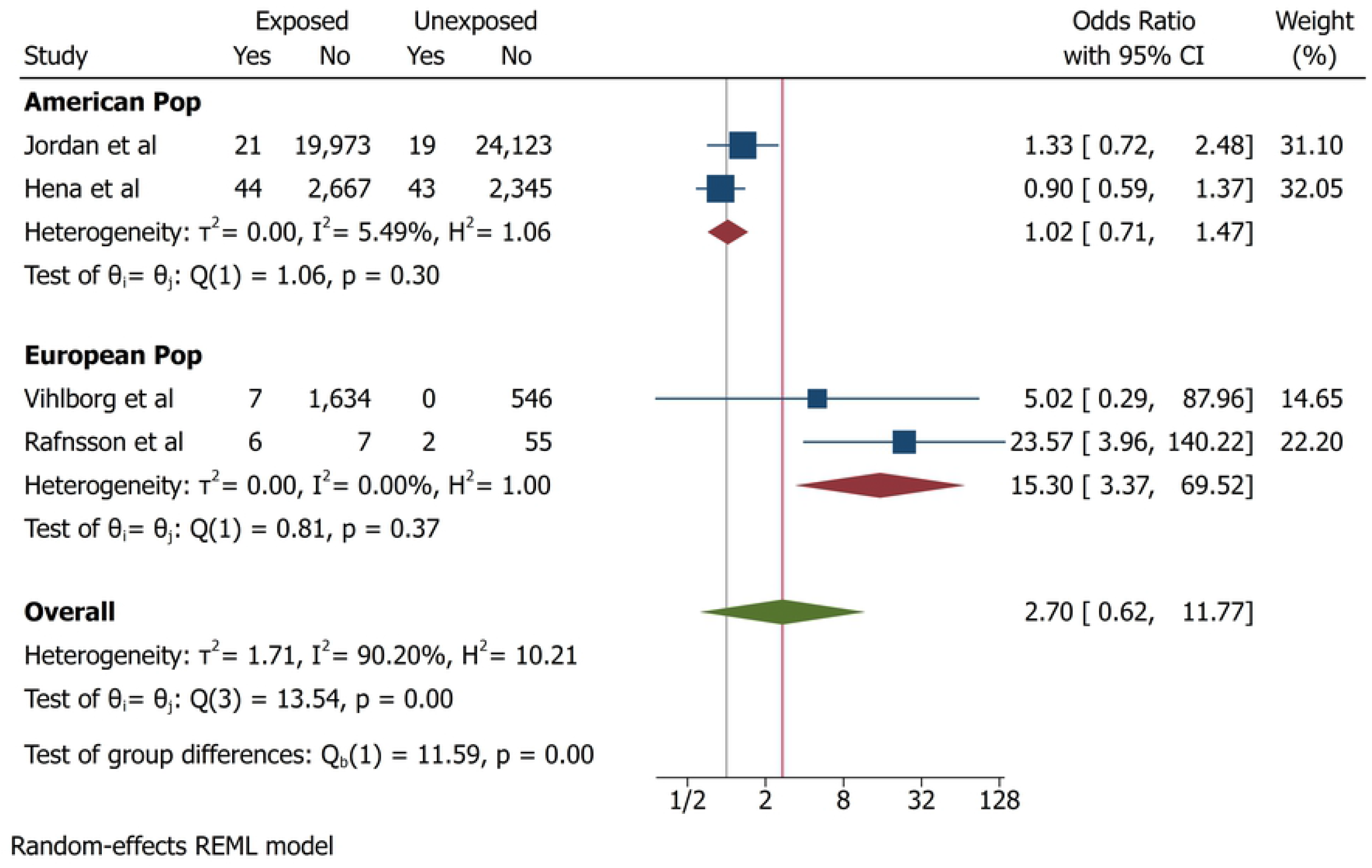
Meta-analysis results of eligible studies (1998-2019)

## Discussion

This study aggregated available evidence from the published literature on exposure to respirable particles and increased risk of developing sarcoidosis. The subgroup analysis in European populations indicated significantly higher risk of sarcoidosis among groups exposed to respirable particles compared to the unexposed groups. However, among the American population, the risk among the exposed was not statistically significant thereby making the overall risk estimate not statistically significant. These discordant results could potentially be explained by differential misclassification of exposure, under ascertainment of sarcoidosis cases or recall decay of exposure in some populations (10). It is also possible that other factors in addition to (or independent of) the occupational/environmental exposures in the Europeans increase the risk of sarcoidosis in that population (3,22). The current study provides qualitative evidence of the risk of developing sarcoidosis from environmental and/or occupational exposure to dust or other forms of PM.

Exposure to respirable PM is known to have adverse health effects especially as it affects the respiratory health of individuals. While the body has mechanisms that eliminate particles in inspired air, some ultrafine particles escape these mechanisms and are deposited onto the respiratory surfaces leading to inflammation and in some cases fibrosis (23). Pathogenicity of respirable particles is dependent on composition, exposure concentration, exposure duration, ventilatory parameters and particle properties such as origin, size, hygroscopicity and solubility (24,25). The smallest particles cause the most damage due to their smaller diameters and large surface area. Several studies have also shown that these small particles can be a direct or indirect source of reactive oxygen species which have been associated with significant inflammation (26–29). Studies among first respondents of 9/11 have linked exposure to dust to the pathogenesis of various pulmonary diseases including sarcoidosis (30–32). Lung and lymph node samples analyzed from patients exposed to dust from the WTC explosion sites revealed presence of PM including silica, aluminum silicates, talc, and titanium dioxide (33)

Over the past several years, epidemiologic and animal model data have suggested that some environmental and occupational exposures put individuals at risk of sarcoidosis and sarcoidosis-like illnesses (36). Some of these exposures occur in occupations such as agriculture, bird handling, metalworking, lifeguards, defense industries, electronic factories, alloy manufacturing industries, mining, transportation, construction, and wood burning (5,33,37–39). Suspected agents from these workplaces include fungal antigens, bacterial antigens, beryllium, aluminum, barium, rare earth metals, silica, carbon nanoparticles, bird protein antigens, inorganic dust, and mold. A study focused on health outcomes of a group of occupants of a water-damaged office building identified an association between the indoor environmental quality and development of respiratory diseases including sarcoidosis (40). In that study, exposure-response relationship between fungal concentrations and respiratory complaints from the occupants was identified. A follow-up study involving occupants who continued to work from the building demonstrated persistent respiratory health problems including hypersensitivity pneumonitis and sarcoidosis (41). In an in vitro study, inflammatory cytokine secretion response was significantly higher in peripheral blood mononuclear cells from patients with pulmonary sarcoidosis compared to healthy subjects following co-stimulation to fungal and bacterial antigens in organic dust (42). Additionally, statistical correlation between high fungal cell wall agents in air samples and secretion of interleukin-12 in sarcoidosis patient cell cultures has been identified (43).

While the exact role played by inorganic particles in the development of sarcoidosis has not been clearly defined, it has been speculated that development of sarcoidosis occurs due to inflammation because of an antigen presented to the CD4+ T lymphocytes. T cells proliferate, differentiate, produce inflammatory cytokines, and attract other cells which results in granuloma formation in the presence of a persistent antigen and an adjuvant signal. Implicated inorganic dusts that could play the role of adjuvants included various silicates, mineral fibers and alkaline dust that was generated in the 2001 collapse of the WTC (36). This systematic review summarized the results from eligible studies that focused on exposure to respirable particles and the development of sarcoidosis. Although the pooled risk estimate in the meta-analysis was not statistically significant, available evidence suggests that individuals exposed to PM have increased disease risk compared to unexposed (4,7,9,10,16,20,44).

This study highlights important aspects of occupational and environmental risk factors that in congruence with other studies underscore their involvement in development of sarcoidosis. The small numbers of studies identified in this review was a limitation in this study. There were only four studies that were utilized in the meta-analysis, which limits the robustness of the pooled risk estimate computed. Also, the few numbers of identified studies did not allow assessment of publication bias risk. Most of the studies synthesized in this review utilized data from the 9/11 WTC first responder’s cohort and therefore may not be generalizable across other populations. More rigorous epidemiologic exposure studies are needed to generate data that would accurately determine the risk and causal pathways through which exposure to respirable particles could lead to the development of sarcoidosis.

## Data Availability

All relevant data are within the manuscript and its Supporting Information files.

